# Epidemiological characterization of asymptomatic carriers of COVID-19 in Colombia

**DOI:** 10.1101/2020.06.18.20134734

**Authors:** Aníbal A. Teherán, Gabriel Camero, Ronald Prado de la Guardia, Carolina Hernández, Giovanny Herrera, Luis M. Pombo, Albert A. Ávila, Carolina Flórez, Esther C. Barros, Luis A. Perez-Garcia, Alberto Paniz-Mondolfi, Juan David Ramírez

**Affiliations:** Red Cross Section Bogotá – Cundinamarca, Colombia; COMPLEXUS Research group, Fundación Universitaria Juan N. Corpas, Colombia; Grupo de Investigaciones Microbiológicas-UR (GIMUR), Departamento de Biología, Facultad de Ciencias Naturales, Universidad del Rosario, Colombia; Grupo de Investigación ANTHUS, Universidad de la Sabana; Instituto Nacional de Salud, Colombia; Instituto de Investigaciones Biomédicas IDB / Incubadora Venezolana de la Ciencia, Cabudare, Edo. Lara, Venezuela; Ichan School of Medicine at Mount Sinai, NY, USA

**Keywords:** COVID-19, Asymptomatic, Carrier States, Risk factors, Novel Coronavirus

## Abstract

**Objective:** Asymptomatic carriers (AC) of the new Severe Acute Respiratory Syndrome Coronavirus 2 (SARS-CoV-2) represent an important source of spread for Coronavirus Disease 2019 (COVID-19). Early diagnosis of these cases is a powerful tool to control the pandemic. Our objective was to characterize patients with AC status and identify associated sociodemographic factors.

**Methods:** Using a cross-sectional design and the national database of daily occurrence of COVID-19, we characterized both socially and demographically all ACs. Additional Correspondence Analysis and Logistic Regression Model were performed to identify characteristics associated with AC state (OR, 95% CI).

**Results:** 2338 ACs (11.8%; 95% CI, 11.3-12.2%) were identified, mainly in epidemiological week 18 [EW] (3.98; 3.24-4.90). Age ≤ 39 years (1.56; 1.42-1.72). Male sex (1.39; 1.26-1.53), cases imported from Argentina, Spain, Peru, Brazil, Costa Rica or Mexico (3.37; 1.47-7.71) and autochthonous cases (4.35; 2.12-8.93) increased the risk of identifying AC. We also identified groups of departments with moderate (3.68; 3.13-4.33) and strong (8.31; 6.10-7.46) association with AC.

**Discussion:** Sociodemographic characteristics strongly associated with AC were identified, which may explain its epidemiological relevance and usefulness to optimize mass screening strategies and prevent person-to-person transmission.

## INTRODUCTION

In March 2nd, 2020, Colombia reported the first case of Coronavirus Disease 2019 (COVID-19), and as of May 23rd, more than 20,000 cases have been confirmed nationwide [1]. Asymptomatic carriers (AC) may be associated with the accelerated growth of cases in the initial phases of the pandemic, inadvertently spreading the infection to close contacts. In this case, transmission can only be limited until a diagnosis of SARS-CoV-2 infection is rendered after (i) isolation due to symptom onset, (ii) contact tracing or (iii) identification during massive screening strategies [2,3].

AC and pre-symptomatic cases are epidemiologically relevant since they represent a silent source of spread in various public settings (e.g. public transportation, emergency rooms, supermarkets, shelters) [4–6]. The proportion of ACs has been estimated at 15-25%, but seroprevalence studies have reported values of up to 43.2% (95% CI, 32.2-54.7%). Nonetheless, many pre-symptomatic patients are wrongfully classified as ACs during the incubation phase; to later become pauci-symptomatic or develop respiratory manifestations ranging from pneumonia to respiratory failure, or exhibit any other clinical symptoms within the COVID-19 spectrum [4–8].

Epidemiological predictive models have been developed and updated to incorporate silent mobility through AC phenotype in anticipation for the second and third epidemic waves of COVID-19. Such is the case for the SEIR model (Susceptible, Exposed, Infected and Recovered), recently updated to SEAIR (Susceptible, Exposed, Asymptomatic, Infected and Recovered) [9]. In China, estimates indicate that 60-65% of ACs remained undetected. Therefore, under the SEIR model and applying machine-learning-based transmission simulators (MLSim), including the number of undetected AC within its parameters and assuming 15 close contacts per day, estimates suggest that as of April 15th, 2020, the United States---the country contributing the majority of cases imported to Colombia---, could have presented 277,641-to-495,128 latent cases of COVID-19, potentially increasing the spread of the virus [10].

The assessment of ACs and the identification of sociodemographic characteristics associated with this subpopulation could be useful to estimate sample calculations in massive screening studies, as well as adjust control and mitigation measures---especially the intensity of isolation. Therefore, the objective of our study was to characterize ACs demographically and socially, as well as to identify individual characteristics in interaction models associated with ACs.

## METHODOLOGY

### Design and patient’s selection

We performed a cross-sectional study with information from the National Institute of Health (INS) database on COVID-19 cases updated until May 23, 2020. By INS protocol, suspected AC cases remained in quarantine for 7 days while monitoring the appearance of symptoms on a daily basis; on the eighth day, a nasal swab sample was collected to identify or rule out AC state. Records without health status information (symptomatic, asymptomatic) were excluded. The database is public, with de-identified patient data and IRB approval was thus exempt.

### Database and variables

We used variables such as date of diagnosis, age, sex, country of origin, department, case type (imported, related), care setting (home, nursing home, hospital, intensive care unit) and outcome (recovered, convalescent, deceased). The date of diagnosis was adjusted into epidemiological weeks (EW), which were later grouped according to the pattern of AC occurrence (Figure S1) in EW 10-15, 16-17, 18, 19-21; additionally, the variable AC [yes, no] was established.

### Statistical analysis

Data are presented in medians or proportions estimated with 95% CI due to the lack of massive screening for COVID-19 in certain areas of the country. The geographical origin and destination of imported cases was represented with a Sankey Plot (SankeyMATIC (BETA). Cumulative trends and case charts were created with the number of daily cases by Epid_weeks (RStudio Version 1.2.5042). In addition, a heatmap analysis was included to depict a dynamic representation of daily cases by Department from March 6th through May 23rd, 2020 (Orange Data Mining & Fruitful Fun, Version 3.25). The proportion of asymptomatic and symptomatic patients and the median age were compared with the Z and U Mann Whitney tests respectively (significant p-value <0.05, two tails) [Addinsoft. 2020. XLSTAT statistical and data analysis solution. New York, USA. https://www.xlstat.com]. Age was dichotomized between 0-39 and ≥ 40 years due to its association with asymptomatic and symptomatic states respectively (preliminary exploratory analysis not shown).

Countries of origin and departments associated with ACs were identified with a Correspondence Analysis (CA). Additionally, with contribution coordinates of the columns (CCC-CA), groups with a variable level of association with ACs were created [Addinsoft. 2020. XLSTAT statistical and data analysis solution. New York, USA. https://www.xlstat.com].

To estimate the association between sociodemographic characteristics with CA (OR 95%),two Logistic Regression Models (LRM) were performed, the first to establish the main effects and the second a step-backward interaction model of the second level (p-value in <0.05; p-value out:> 0.1), which used the lowest Akaike criteria to select the best model (JASP Team (2020). JASP (Version 0.12.2) [Computer software]).

## RESULTS

### General characteristics

We identified 2,388 ACs (11.8%; 11.3-12.2%) out of 20,177 cases reported in the database. Four cases were excluded due to lack of health status information. The occurrence of AC state in relation to symptomatic presented a slow growth phase between EW 10-15, moderate growth between EW 16-17, and a peak at EW 18, followed by a decrease between EW 19-21 (Figure 1a, 1b, S1). Daily cases ranged from 1 to 203 per day, and Ews 18 and 19 registered the highest number of cases per day: 172 and 203, respectively.

**Figure 1.**
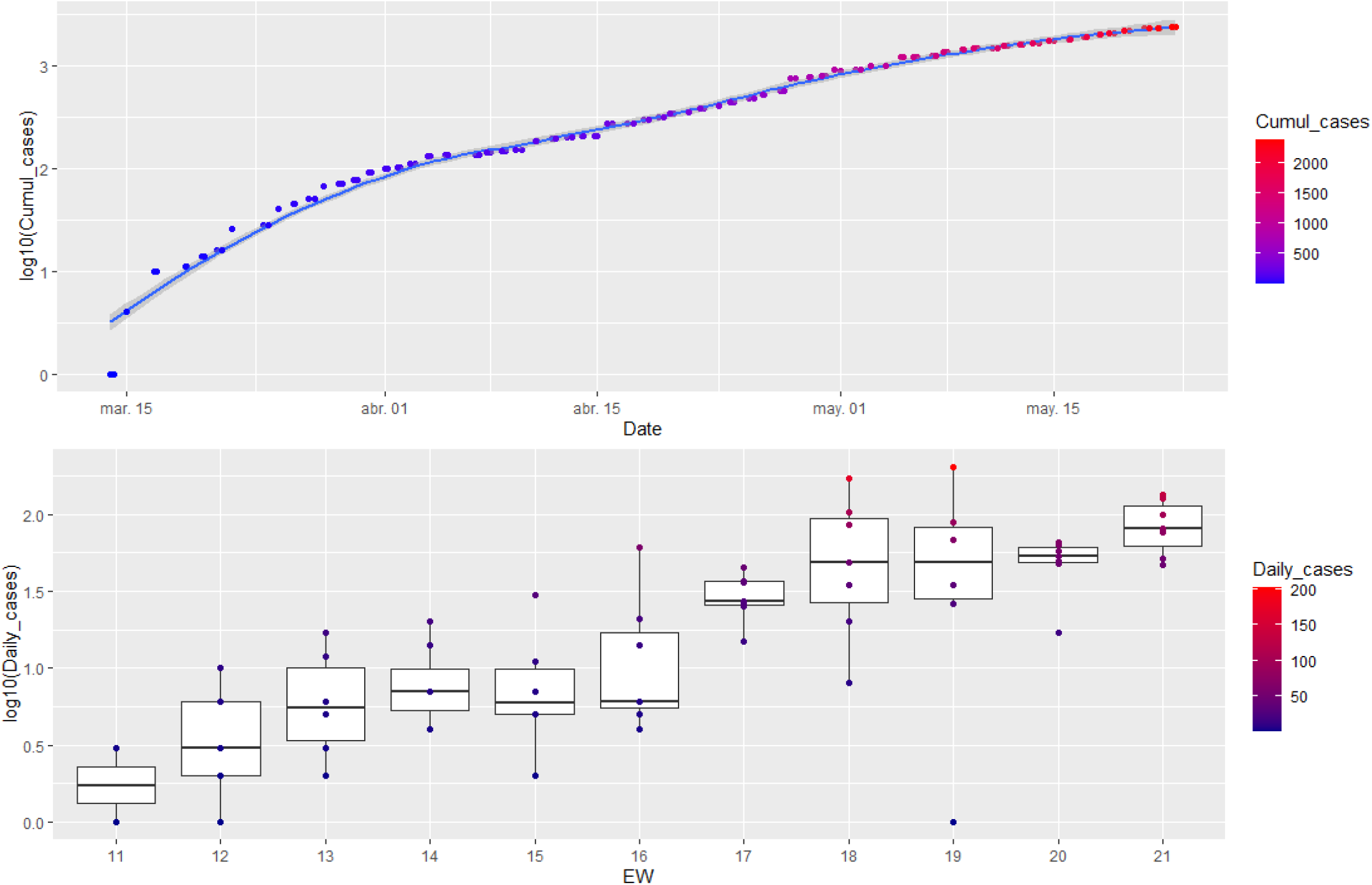
Daily accumulation and distribution of ACs by epidemiological week in Colombia. The y-axis represents the number of daily ACs transformed into a base 10 logarithm. In the bottom figure, each point represents the number of daily cases in each epidemiological week (EW). In both figures, the number of cases per day is located in points that increase in color intensity according to the occurrence of cases.

Additionally, we report department clusters with a high occurrence of daily COVID-19 cases, which follow different dynamic patterns for ACs and symptomatic patients (Figure 2). Meta reported the largest number of daily ACs (n: 151), with peak occurrences between April 23rd and May 10th; followed by Amazonas, Bogotá, and Caribbean departments with peak reports between May 11th and 12th,

**Figure 2.**
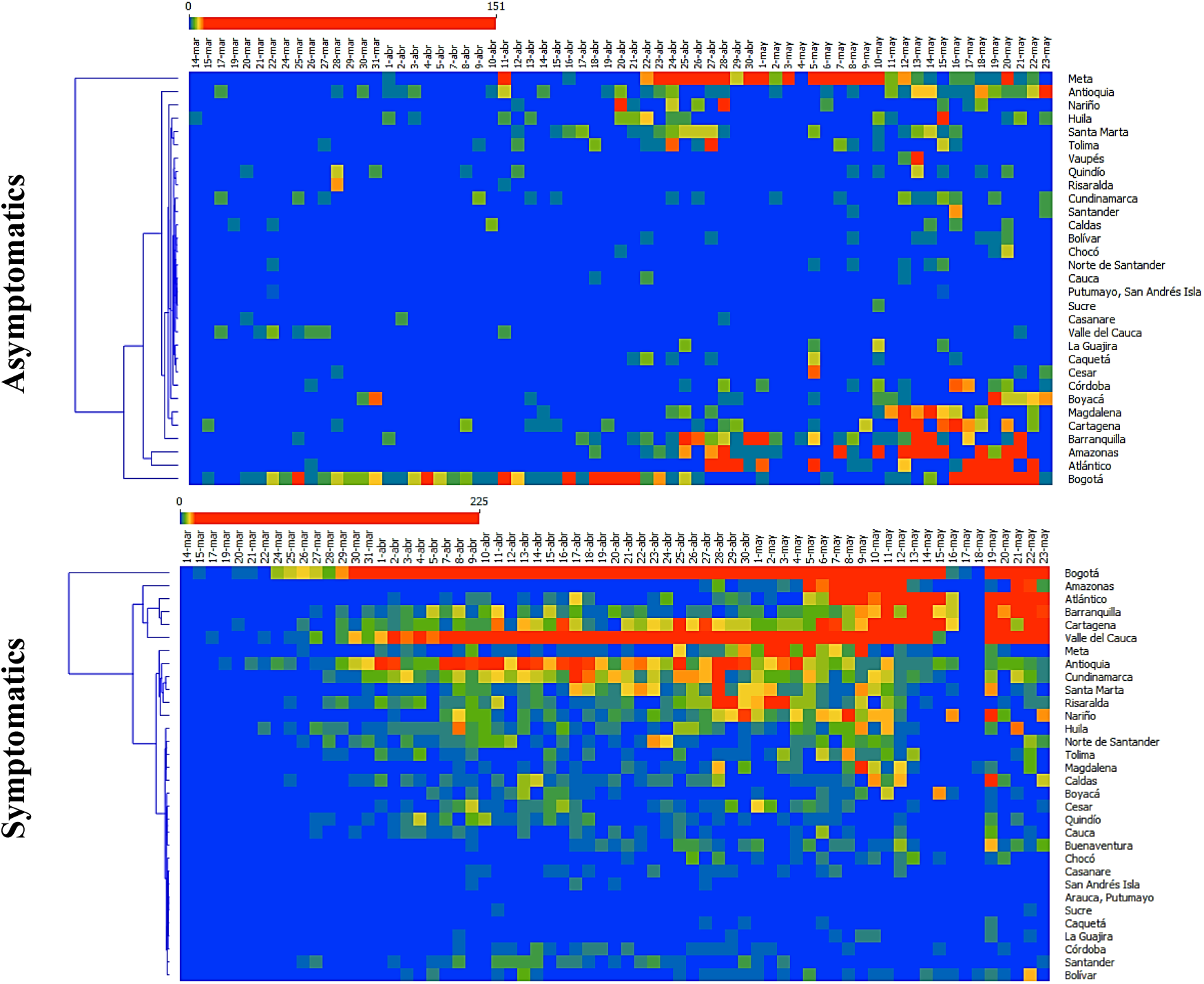
Dynamics of daily reports of COVID-19 in Colombia by Department. Heatmap showcasing the number of ACs (top) and symptomatic patients (bottom) diagnosed in every Colombian department until May 23rd, 2020.

More than half of the imported ACs came from Europe, specifically Spain, followed by North and South America. Those that arrived from Spain and USA were distributed mainly in Bogotá, Valle del Cauca and other departments of the Caribbean region. Amazonas department only received imported ACs from South American countries. The origin and distribution of imported symptomatic patients was more diverse; however, most cases originated from Spain, USA, Ecuador, Mexico, Brazil, or Panama, and were mainly distributed across Bogotá, Antioquia, and Valle del Cauca (Figure 3b and 3b).

**Figure 3.**
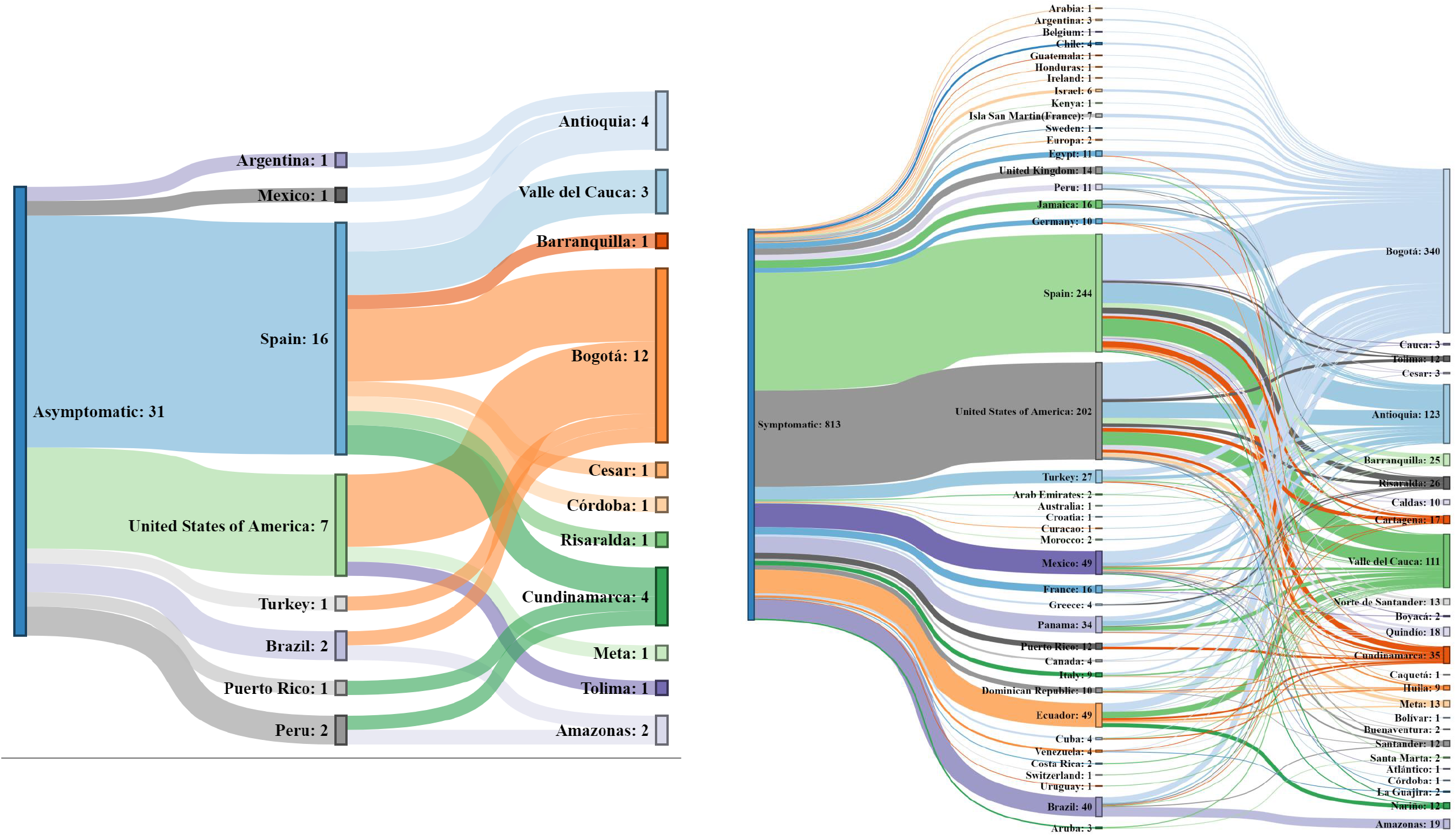
Origin and destination of imported asymptomatic and symptomatic cases. The left and right figures, respectively, represent the country of origin and destination department of ACs and symptomatic patients. The thickness of the link tapes corresponds to the number of reported cases.

About half of the ACs were located in Meta and Bogotá, and a tenth in the Amazon (Table 1, Table S1). Median age was 32 years old, lower than the symptomatic patients. Most of them were males (Table 1). The domicile was the main place of care (85.7%; 84.3-87.1%) and 13.9% had recovered (95%, CI, 12.5-15.3%). Four of the admitted cases had fatal outcomes, two from the general wards and two from the Intensive Care Units (ICU). Possibly these last 8 cases were treated for symptoms unrelated to COVID-19, or were diagnosed post-mortem.

**Table 1.** Sociodemographic characteristics of the asymptomatic and symptomatic patients

| **Variables** | **Asymptomatic** | **Symptomatic** | **p-value** |
| --- | --- | --- | --- |
|  | **n: 2388** | **n: 17785** |  |
| **Age, years** | 32 (24-44) | 37 (26-53) | <0.001 |
| **0-39** | 1603 (67.1) | 9655 (54.2) | <0.001 |
| **≥40** | 785 (32.8) | 8130 (45.7) | <0.001 |
| **Sex** |  |  |  |
| **Male** | 1208 (75.2) | 6220 (53.7) | <0.001 |
| **Female** | 399 (24.8) | 5359 (46.3) | <0.001 |
| **Geographical source** |  |  |  |
| **Related** | 2357 (98.7) | 16972 (95.4) | <0.001 |
| **Imported** | 31 (1.30) | 813 (4.57) | <0.001 |
| **Departments†** |  |  |  |
| **Meta** | 749 (31.3) | 219 (1.23) | <0.001 |
| **Bogotá** | 437 (18.3) | 65.4 (36.8) | <0.001 |
| **Amazonas** | 238 (9.97) | 1150 (6.47) | <0.001 |
| **Atlántico** | 203 (8.50) | 1068 (6.01) | <0.001 |
| **Barranquilla** | 117 (4.90) | 1216 (6.84) | <0.001 |
| **Imported cases** |  |  |  |
| **Spain** | 16 (51.6) | 244 (30.0) | 0.011 |
| **USA** | 7 (22.5) | 202 (24.8) | 0.774 |
| **Brazil** | 2 (6.45) | 40 (4.92) | 0.700 |
| **Mexico** | 2 (6.45) | 49 (6.03) | 0.922 |
| **Argentina** | 1 (3.23) | 3 (0.37) | 0.023 |
| **Peru** | 1 (3.23) | 11 (1.35) | 0.387 |
| **Puerto Rico** | 1 (3.23) | 12 (1.48) | 0.437 |
| **Turkey** | 1 (3.23) | 27 (3.32) | 0.977 |

†: cases that appeared spontaneously in Colombia, ††: top 5 of the departments with the highest frequency of AC.

### Factors associated with AC condition

Using the CCC-CA, a group of six countries and three groups of departments were associated with AC state (Figure 4). To execute LRMs, the variables “age group 0-39 years” and “male sex” were transformed into dummi (0/1). With a preliminary LRM, a higher β coefficient was estimated in relation to cases imported from countries associated with AC, therefore, the variable “geographical origin” was created, composed of the categories “imported from countries associated with symptomatic” [Imported CAS - referent], “imported from countries associated with asymptomatic” [Imported CA-AC] and “related cases”. Additionally, a variable was created for the departments grouped with the CA [departments with low association - referent] and for the EW (EW 10-15 - referent). The first LRM (main effects) identified a significant association of all index sociodemographic categories with AC state (Table S2). The second model explores the following interactions: 1. Geographical origin and grouped departments, 2. Geographical origin and EW, 3. Age group (0-39 years) and gender, 4. Grouped departments and gender; and, 5. Age (0-39 years) and EW. We identified that the variables gender (males) and EW showed interaction with the grouped departments (Table 2).

**Table 2.** Factors associated with asymptomatic carrier (AC) state in Colombia

| Variable | Interaction model |  |  |
| --- | --- | --- | --- |
|  | B | ORc (95%, CI) | p-value |
| **Intercept** | **-4.531** | - | **<0.001** |
| **Age** |  |  |  |
| >40 years | **Ref.** | - | - |
| 0-39 years | 0.451 | 1.569 (1.422-1.732) | **<0.001** |
| **Sex** |  |  |  |
| Female | **Ref.** | - | - |
| Male | -0.019 | 0.981 (0.850-1.132) | 0.791 |
| **Department** |  |  |  |
| Low association [1] | **Ref.** | - | - |
| Moderate association [2] | 0.210 | 1.234 (0.799-1.908) | 0.344 |
| Strong association [3] | 1.130 | 3.095 (1.860-5.149) | **<0.001** |
| **Geographical source** |  |  |  |
| Imported CAS<sup>†</sup> | **Ref.** | - | - |
| Imported CA-AC<sup>††</sup> | 1.225 | 3.405 (1.491-7.775) | **<0.004** |
| Related cases<sup>†††</sup> | 1.351 | 4.861 (1.885-7.910) | **<0.001** |
| **EW** |  |  |  |
| 10-15 [1] | **Ref.** | - |  |
| 16-17 [2] | 0.657 | 1.410 (1.155-1.721) | **<0.001** |
| 18 [4] | 0.418 | 1.519 (1.087-2.122) | **<0.014** |
| 19-21 [3] | -0.074 | 0.929 (0.758-1.138) | 0.477 |
| **Department [2] + Male** | 0.363 | 1.438 (1.035-1.998) | **0.030** |
| **Department [3] + Male** | 0.757 | 2.132 (1.723-2.638) | **<0.001** |
| **EW [2] + Department [3]** | -1.052 | 0.349 (0.198-0.616) | **<0.001** |
| **EW [3] + Department [2]** | 1.474 | 4.368 (2.780-6.862) | **<0.001** |
| **EW [4] + Department [2]** | 1.219 | 3.385 (1.631-7.023) | **0.001** |
| **EW [4] + Department [3]** | 1.414 | 4.673 (2.273-7.447) | **<0.001** |

†: CAS countries associated with symptomatic patients, ††: CA-AC countries associated with asymptomatic carriers (AC), †††: spontaneous cases, **EW**: epidemiological weeks.

**Figure 4.**
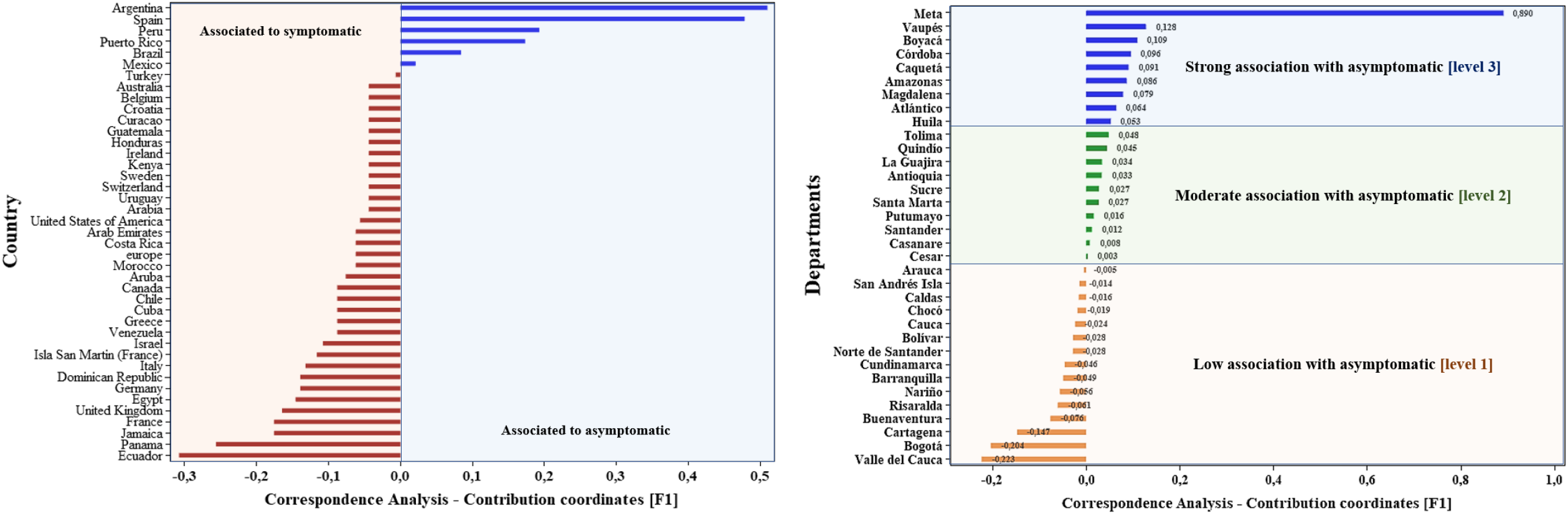
Groups of countries and departments associated with AC state. The left figure shows the group of countries associated with asymptomatic carrier (AC) state identified with positive values of the CCC-CA. The right figure shows departments grouped according to three intervals of CCC-CA: low association (CCC-CA: negative values), moderate association (CCC-CA:> 0 - <0.05), and strong association (CCC-CA: ≥ 0.5).

Variables “0-39 years”, “departments with strong association”, “imported CA-AC” and “related cases” were found to increase the risk of identifying AC state. It was also determined that the risk increased with the interaction between men in the departments with a strong and moderate association. In isolation (without interaction), between EWs 19-21, the risk of identifying AC decreased, as did EWs 16-17 in departments with a strong association. However, the risk increased from EW 18 to EWs 19-21 when interacting, both with moderate and strong association departments (Table 2).

## DISCUSSION

We found that, in an isolated fashion, age < 40 years old, imported cases from a group of 6 countries, autochthonous cases and the occurrence in groupings of departments were associated with AC state. Additionally, the risk of being a male AC was only identified in departments with moderate or strong risk, and the risk was variable in the groupings of departments throughout specific epidemiological periods.

Additionally, our results show that the proportion of ACs in Colombia lays between 11-12% (Table 1), a lower estimate than previously described in other case series or mass screening studies with reported proportions between 5-80% [11–14]. Given the inclusion of presymptomatic patients or the unification of AC with non-critical symptoms in some reports, we can’t rule out that a non-differential classification bias influenced these estimates. An adapted definition for AC in Colombia may address this limitation.

Figure 1 shows that the majority of imported cases to Colombia came from Spain and USA, where AC rates have been estimated at 2.5% and 25%, respectively [14,15]. Although imported cases carry a distinctive genetic load that, population-wise, could manifest itself as a particular phenotype [16], currently there are no reports of genetic variants associated with AC in general or for any of the four AC subtypes described in the literature [17]. Subsequent research should be conducted on the possible association between ACs and phylogenetic variants (or other variables) to support the differential risk identified in imported cases from different regions of the world.

We identified that imported cases from a group of 6 countries were strongly associated with AC (Figure 3, Table S1, Table 2), and although no interaction was established between the country of import and the destination department (data not shown), we observed that departments strongly associated with AC had less diversity of import origin. Such is the case of Meta and Amazonas, which exclusively imported cases from USA and Brazil/Peru, respectively (Figure 2).

Among the demographic characteristics, the association between AC state and patients under 40 years of age stands out. Possible explanations for this observation include: (i) the lower presence of co-morbid conditions and baseline health issues within this age group and (ii) the higher risk of exposure through work activities which are greater in this age group [18]. However, clinical or social environment could also explain this finding, as a study in skilled nursing facility residents showed a high proportion of AC in those over 70 years of age, however this was a premature finding since most patients were later reclassified as pre-symptomatic or pauci-symptomatic [11].

We identified a higher frequency of men infected with COVID-19 consistent with reports from other countries around the world, except in Spain and Switzerland, where women ranked first [19]. Frequent occupations performed by men, as well as certain immunological and genetically susceptible backgrounds have been associated with this finding [19,20]. In particular, the risk of being an AC was higher in men, and increased in geographic areas associated with AC. This interaction is not uncommon given that professions regularly carried out by men, including those such as taxi driving, private security or prison guarding, among other work settings, can be distributed asymmetrically within countries, a pattern that would explain our findings [20].

The phases on the occurrence of cases throughout EWs and the interaction with groupings within departments associated with AC has been previously described in Chongqing, China, where researchers identified significant changes in the frequency of cases after implementation of geographic isolation measures. In Wuhan, a study showed that one group of ACs was linked to imported cases while others were linked mostly to autochthonous cases from geographically isolated areas of Wuhan [21]. We identified that in addition to being associated with a travel history to foreign countries, ACs were also associated with cases that appear spontaneously (related), occurring differentially as measures of geographic and social isolation were applied.

The lack of mass screening for COVID-19 in Colombia is the main limitation of our study since the actual AC ratio and the distribution of specific characteristics may differ from those estimated in this report. On the other hand, although a cross-sectional design is not ideal to identify risk factors, to the best of our knowledge this is the first study aimed at identifying factors associated with AC state with population data unbiased by the inclusion of pre-symptomatic cases.

The COVID-19 pandemic has had serious socioeconomic implications, including a collapse of healthcare systems, bankruptcy of companies as well as increasing trends in unemployment and crime rates [22–25]. This has forced countries with limited resources---such as Colombia---to perform massive screenings in order to prematurely lift quarantine and isolation measures despite the latent risk of successive outbreaks caused by a potential silent spread of COVID-19 through cases in the pre-symptomatic phase or AC state [26,27].

ACs transmit COVID-19 more efficiently than symptomatic patients for up to 21 days after the presumed date of infection [28,29]. This led to their inclusion in mathematical models intended to estimate the probability or expected number of person-to-person infections on repatriation trips from Wuhan, China [7,30]. Since then, ACs have become the target of mass screening in Asian and European countries effectively reducing economical losses due to unnecessary hospital care, controlling the spread in public or in-hospital settings, and allowing the execution of safe plans of social and work re-integration after quarantine and isolation [26,31–35].

To date, testing of asymptomatic individuals rests at the discretion of physicians when justified on a case-by-case basis. On the other hand, the utility of SARS-CoV-2 testing for broad screening of asymptomatic individuals remains to be determined given the limited sensitivity data available for most commercially available test kits [36].

Together, our findings demonstrate sociodemographic trends strongly associated with COVID-19 AC state in Colombia at a departmental and national level. We believe that the implementation of massive screening campaigns to detect AC and pre-symptomatic patients is paramount to further characterize this phenomenon and adequately guide public health measures of containment and prevention. Additional molecular analysis of viral and host genotypic characteristics should be conducted to determine possible associations with AC state.

## Data Availability

The data are in their relevant supporting information

## Author Contributions

All authors have read and agreed to the published version of the manuscript.

